# Demographic and professional risk factors of COVID-19 infections among physicians in low- and middle-income settings; findings from a representative survey in two Brazilian states

**DOI:** 10.1101/2022.05.27.22275696

**Authors:** Giuliano Russo, Alex Cassenote, Bruno Luciano Carneiro Alves de Oliveira, Mario Scheffer

## Abstract

**Introduction:** Health workers (HWs) are a key resource for health systems worldwide, and have been affected heavily by the COVID-19 pandemic. Evidence is consolidating on incidence and associated drivers of infections, predominantly in high-income settings. It is however still unclear what the risk factors may be for specific health professions, particularly in low- and middle-income countries (LMICs).

**Methods:** We conducted a cross-sectional survey in a representative sample of 1,183 medical doctors registered with Brazil’s Federal Council of Medicine in one developed (São Paulo) and one disadvantaged state (Maranhão). Between February-June 2021, we administered a telephone questionnaire to collect data on physicians’ demographics, deployment to services, vaccination status, and self-reported COVID-19 infections. We performed descriptive, univariate, and multilevel clustered analysis to explore the association between physicians’ infection rates, and their sociodemographic and employment characteristics. A generalized linear mixed model with a binomial distribution was used to estimate the adjusted odds ratio.

**Results:** We found that 35.8% of physicians in our sample were infected with COVID-19 in the first year of the pandemic. The infection rate in Maranhão (49.2%) [95% CI 45.0-53.4] was almost twice that in São Paulo (24.1%) [95% CI 20.8-27.5]. Being a physician in Maranhão [95% CI 2.08-3.57], younger than 50 years [95% CI 1.41-2.89], and having worked in a COVID-19 ward [95% CI 1.28-2.27], were positively associated with the probability of infections. Conversely, working with diagnostic services [95% CI 0.53-0.96], in administrative functions [95% CI 0.42-0.80], or in teaching and research [95% CI 0.48-0.91] was negatively associated.

**Conclusions:** Based on data from Brazil, COVID-19 infections in LMICs may be more likely in those health systems with lower physician-to-patient ratios, and younger doctors working in COVID-19 wards may be infected more frequently. Such findings may be used to identify policies to mitigate COVID-19 effects on HWs in LMICs.

## 1 Introduction

Health workers (HWs) are a crucial healthcare resource, as they deliver services, manage the health system’s building blocks, and represent the single most important health expenditure across the world.(1) Their role and importance have been particularly evident during COVID-19, when they have been at the frontline of curative and preventive services, and led the clinical response to the pandemic.(2) Precisely because of their exposure, many HWs across the world contracted the virus, and some have died.(3)

It is not clear how many HWs have been lost to COVID-19, and what the key risk-factors may be. The World Health Organization estimates that between 80,000 and 180,000 may have actually died, if country-specific COVID-19 infection and fatality rates were to be applied to the 135 million-strong global health workforce (4). It is unclear which health professions were more affected, and what the specific risk factors could be, particularly for low- and middle-income countries (LMICs).(5)

Cross-sectional, cohort, and hospital-based prevalence studies have been carried out to estimate COVID-19 infections among health workers. An observational cohort study from Denmark (6) screening for COVID-19 infections among medical, nursing and students personnel identified 4·04% seropositive health-care workers. A metanalysis of the prevalence of staff infections in hospital settings from 47 eligible studies in America, Europe and Asia,(7) found the prevalence of infection was 7% for those studies using antibody tests, while for those studies using PCR tests, prevalence of infections was 11%. A highly referenced systematic review of the evidence from the first semester of the pandemic(8) estimated a total of 152,888 infections among health workers and 1,413 deaths worldwide.

A living review of the epidemiology of COVID-19 among HWs (9) concluded that these account for a significant proportion of global coronavirus infections worldwide; nurses may be the personnel most at risk, and illness severity is lower in non-patient facing workers.(10) Professional exposures such as involvement in intubations, direct patient contact, or contact with bodily secretions, were associated with increased infection risk. However, there is evidence that private community exposure may be a stronger risk factor than work exposure.(11)

Exposure to at-risk-patients from high-infection regions or from dedicated COVID-19 wards, would be relevant risk factors,(12) but it is not clear what specific healthcare settings would increase the risk of infection for HWs, and who exactly are the more exposed ‘frontline workers’. A study assessing COVID-19 hospital admission in Scotland(13) found that older, male HWs were more at risk of infection and hospitalization; patient-facing healthcare workers and their households were at higher risk compared with non-patient facing ones. A prospective, observational cohort study in the UK and the US (14) concluded that self-reported infection rates among frontline health-care workers are higher than in the general population. On the other hand, a study from a UK hospital(15) showed that infection levels were greatest among HWs working in housekeeping, acute medicine, and general internal medicine, with surprisingly lower rates for those working in intensive care.

Less evidence is available from LMICs. An analysis of 101 medical staff admitted for COVID-19 in a Wuhan hospital in China (16) showed these to be younger than typical patients, and displaying milder symptoms. A study from Iran(17) also found COVID-19 infections among HWs to be greater among younger (<35) professionals, particularly among women and nurses. Another study in 14 hospitals from Qatar(18) found HWs infections to be more frequent in non-COVID-19 facilities, where Personal Protective Equipment (PPE) would be only erratically used. Another investigation of COVID-19 cases among HWs in Oman(19) confirmed that the majority of these were among young (>45 years), female workers, predominantly deployed in primary care settings.

A study assessing COVID-19 infections among health workers in a Rio de Janeiro hospital in Brazil(20) found an overall seroprevalence of 30%. Non-white staff (mostly hospital support workers) with lower income and schooling, as well as users of the mass transportation system, showed the highest infection rates. HWs income level, schooling and work modality appeared as negative predictors. Analogous studies from São Paulo hospitals(21)(22) showed HWs infection rates to be similar to those in the wider population. Male and non-clinical workers appeared to be more at risk but working in COVID-19 services was not associated with higher levels of infections.

A knowledge gap exists on the association of HWs personal and professional characteristics with COVID-19 infections in low-income settings, particularly for doctors. This is particularly of interest in South America which has been hit particularly hard by the epidemic(23); an estimated 13,525 health workers died in Brazil, the world’s second largest loss.(4)

In this paper we report the results of a study to explore the self-reported COVID-19 infection rates among physicians in Brazil, with a view to identify the risk factors associated with infections. The study aimed at contributing to the existing knowledge on impacts of COVID-19 on health workers in LMICs, providing an evidence base for local and international policies to mitigate effects of the pandemic.

## 2 Methods

As part of a research project on the impact of COVID-19 and the associated economic crisis on Brazil’s health system,(24) we conducted a representative cross-sectional telephone survey among registered physician during the second year of the pandemic, in two states, São Paulo and Maranhão.

These represent extreme cases of economic and health system disparities.(25) With almost 47 million people, São Paulo has one of Brazil’s highest income per capita (US$ 12,776), and 43% of its population is covered by private health insurance schemes.(26) Conversely, Maranhão has slightly more than 7 million people, its income is about one third of São Paulo’s, and just 7% of the population own a private health plan (Tab.1).

São Paulo has three times the number of physicians per capita as Maranhão.(27) São Paulo appears to have been hit harder by the pandemic,(28) with three times the number of COVID-19 deaths per capita at the time of our study, and twice the number of infections among its population (Tab.1).

### 2.1 Data collection and sampling strategy

The physician database for the two states was provided by Brazil’s Federal Council of Medicine; the survey sample was calculated by the Faculty of Medicine of the University of São Paulo, and the actual survey was carried out by the survey services institute ‘*Datafolha’*, under the technical supervision of the academic partners of the study. This survey is part of a wider study on the impact of COVID-19 and economic recessions on Brazil’s health system and workforce.(24)

A representative cross-sectional study including 1,183 physicians was conducted in 2021. Two independent sample sizes per state was calculated based on a total of 152,511 active medical registries in São Paulo (N=144,852) and Maranhão (N=7,659) from the Federal Medical Council Medicine database (*Conselho Federal de Medicina - CFM*), using a 95% confidence level with 5% margin of error and statistical power of 80% (see the two Simple Random Sampling with Replacement equations in Supporting information 1). Proportional stratified sampling was drawn distribution for gender, age, state and local of address (capital or countryside) were preserved.

Substitution was carried out exclusively in cases of unsuccessful contact or refusal to participate in our survey; 1,183 physicians were randomly selected, and five substitutions were identified for each sampled physician. Substitution sampling followed the same stratification criteria used for the initial sample calculation. We controlled sample replacements for state, sex, and age, so that every physician who did not agree to participate was replaced by an individual with the same gender, age, and specialty characteristics to avoid selection bias.

Primary data were collected via a telephone survey carried out by 8 data collectors, including one field coordinator, 6 experienced interviewers, and two administrative staff responsible for checking missing data. Sample size calculations, sample selection, questionnaire design, substitution control, database assembly and data analysis were performed by the authors of this papers. Three senior researchers from the medical demography field previously piloted and calibrated the questionnaire with 30 interviewees to estimate the substituion rate. Reproducibility was tested by sampling a random sample after the field collection and repeating the interview, resulting in 100% agreement.

Data collection was carried out between the 16^th^ February and 15^th^June 2021 by the Datafolha Research Institute under supervision of the authors’ research institutions. The interviews consisted of a 30-minute telephone questionnaire, containing 30 questions ranging from multiple, closed questions to interdependently concatenated and semi-opened questions (see Supporting information 2: Survey Questionnaire).

Patient and public involvement. Medical doctors as well as members of the public were consulted and participated in the design of the original version of the survey questionnaire. The questionnaire was then piloted by Datafolha in a subset of ten doctors in the two states, and a final version was elaborated following the feedback received.

### 2.2 Data analysis

Self-reported COVID-19 infection events during the past year was the selected outcome variable, distinguishing between the severity of the infection events (asymptomatic, mild, and severe). We used sociodemographic characteristics (sex, gender, age, and income), medical employment characteristics (such as medical specialty, training and years of service), and type of work carried out in COVID-19 and regular wards (intensive care, inpatient or outpatient care, distance-based consultation, or research), as independent, explanatory variables.

We performed descriptive, univariate, and multilevel clustered analysis on the dataset. A generalized linear mixed model with a binomial distribution was used to estimate the adjusted odds ratio(29) as these allow for the inclusion of random or cluster-specific effects in the linear predictor. The inclusion of random effects in the linear predictor reflects the idea that there is natural heterogeneity across geographic clusters in their regression coefficients. Such method has been used extensively in health services research.(30) The ‘enter’ method was used for the selection of variables, and ANOVA tests were employed to verify the equality hypothesis among the different models. Data were shown as absolute frequency and proportion with a 95% confidence interval. The adjustment of different models was verified by indicators of residual deviance and the Akaike information criterion (AIC).(31) The database developed by the Datafolha data collectors was exported to the Statistical Package for the Social Sciences (SPSS) version 26 for Windows (International Business Machines Corp, New York, USA) and R-GUI version 3.5.3(32) for statistical analysis. All the significance levels were set to p < 0.05.

### 2.3 Ethics considerations

This study received the approval from the Research Ethics Committees of the Federal University of Maranhão (CEP UFMA 3.051.875), and from the Faculty of Medicine of the University of São Paulo (CEP FMUSP 3.136.269), and was approved by Brazil’s Federal Council of Medicine, that provided the list of physicians registered with the council of medicines in the two states, and the respective telephone contact details.

All the physicians contacted via telephone were informed beforehand of the objectives of the survey, and their consent was verbally obtained to participate in thesurvey. All the forms and questionnaires were anonymized through a double coding system organized by the Faculty of Medicine of the University of São Paulo.

## 3 Results

Our sample included 1,183 physicians, 551 from Maranhão and 632 from São Paulo, approximately split equally between urban and rural areas (Table 2 below). Most physicians in our survey (61.6%) worked both in public and private sector jobs (dual practice), with only 25.4% of them employed exclusively in the public. Most of our physicians declared engaging in outpatient care (82.7%) in hospital or clinics settings, and 63.4% of them were deployed directly to the delivery of COVID-19 services, to COVID-19 wards or to COVID-19-specific outpatient care. Engagement in the delivery of specific healthcare services was non-exclusive, as many physicians declared to work concomitantly in multiple wards, services, and sectors.

**Table 1:** Selected economic, health system, and COVID-19 indicators for the study locations at the time of the study.

| Area | Population (2021)* | GDP per capita (USD 2019)* | Phys/100,000 ** | COVID-19 Cases/100,000*** | COVID-19 Deaths/100,000 *** | COVID-19 fatality*** |
| --- | --- | --- | --- | --- | --- | --- |
| Brazil | 213,317,639 | 8,924 | 251.3 | 12,815.6 | 298.2 | 2.2% |
| São Paulo | 46,649,132 | 12,776 | 310.5 | 9,345.2 | 319.1 | 3.4% |
| Maranhão | 7,153,262 | 3,453 | 107.1 | 5,030.3 | 143.4 | 2.9% |

Source: \*IBGE, 2022; \*\* Brazilian Medical Council, 2021; \*\*\* CONASS, 2021

**Table 2:** Characteristics of physicians in the survey sample.

| Characteristics/variables | n | Proportions and Confidence Intervals |
| --- | --- | --- |
| Gender |  |  |
| Male | 665 | 56.2% (53.4%-59.0%) |
| Female | 518 | 43.8% (41.0%-46.6%) |
| Age |  |  |
| < 35 yeas | 404 | 34.2% (31.5%-36.9%) |
| 35 to 50 years | 405 | 34.2% (31.6%-37.0%) |
| > 50 years | 374 | 31.6% (29.0%-34.3%) |
| State |  |  |
| Maranhão (MA) | 551 | 46.6% (43.7%-49.4%) |
| São Paulo (SP) | 632 | 53.4% (50.6%-56.3%) |
| Geographical location of deployment |  |  |
| Rural areas (Interior) | 598 | 50.5% (47.7%-53.4%) |
| Urban areas around capital cities | 585 | 49.5% (46.6%-52.3%) |
| Health sector of deployment |  |  |
| Exclusively Private | 153 | 12.9% (11.1%-14.9%) |
| Exclusively Public | 301 | 25.4% (23.0%-28.0%) |
| Dual practice | 729 | 61.6% (58.8%-64.4%) |
| Employment in specific health services |  |  |
| Outpatient clinical services (hospital or clinics) | 978 | 82.7% (80.4%-84.7%) |
| Diagnostic tests equipment-related services | 392 | 33.1% (30.5%-35.9%) |
| Surgery (in-patient care) | 459 | 38.8% (36.1%-41.6%) |
| Outpatient surgery | 450 | 38.0% (35.3%-40.8%) |
| Administrative position | 288 | 24.3% (22.0%-26.9%) |
| Teaching and research | 312 | 26.4% (23.9%-28.9%) |
| Engagement with COVID-19 services |  |  |
| No | 433 | 36.6% (33.9%-39.4%) |
| Currently working with COVID-19 services | 631 | 53.3% (50.5%-56.2%) |
| Worked in the past, but not currently | 119 | 10.1% (8.4%-11.9%) |
| Type of COVID-19 services delivered |  |  |
| COVID-19 ward or Intensive care unit (ICU) | 524 | 44.3% (41.5%-47.2%) |
| Outpatient COVID-19 care | 490 | 41.5% (38.7%-44.3%) |
| Telemedicine or other distance-based COVID-19 services | 178 | 15.1% (13.1%-17.2%) |
| Research on COVID-19 | 61 | 5.2% (4.0%-6.5%) |
| Epidemiological surveillance or COVID-19 boards | 53 | 4.5% (3.4%-5.8%) |

**Table 3:** Multivariate regression model for physicians personal and professional characteristics and probability of being infected with COVID-19.

|  | B | S.E. | OR (95%CI) | p-value |
| --- | --- | --- | --- | --- |
| Global Model (n=1,183) |  |  |  |  |
| Maranhão state (n=551) | 1.004 | 0.138 | 2.72(2.08-3.57) | <0.001 |
| Gender Female (n=518) | -0.117 | 0.138 | 0.88(0.67-1.16) | 0.396 |
| Age |  |  |  |  |
| > 50 years (n=374) |  |  | Ref. | Ref. |
| 35 to 50 years (n=405) | 0.706 | 0.182 | 2.02(1.41-2.89) | <0.001 |
| < 35 years (n=404) | 0.577 | 0.173 | 1.78(1.27-2.50) | 0.001 |
| Employment in specific health services |  |  |  |  |
| Outpatient clinical services (hospital or clinics) (n=978) | -0.289 | 0.176 | 0.74(0.53-1.05) | 0.102 |
| Diagnostic tests equipment-related services (n=392) | -0.330 | 0.147 | 0.71(0.53-0.96) | 0.025 |
| Surgery (in-patient care) (n=459) | 0.286 | 0.155 | 1.33(0.98-1.80) | 0.065 |
| Outpatient surgery (n=450) | 0.047 | 0.155 | 1.04(0.77-1.42) | 0.760 |
| Administrative position (n=288) | -0.533 | 0.160 | 0.58(0.42-0.80) | 0.001 |
| Teaching and research (n=312) | -0.407 | 0.162 | 0.66(0.48-0.91) | 0.012 |
| Health sector |  |  |  |  |
| Exclusively Private (n=153) |  |  | Ref. | Ref. |
| Exclusively Public (n=301) | 0.248 | 0.256 | 1.28(0.77-2.11) | 0.333 |
| Dual practice (n=729) | 0.129 | 0.236 | 1.13(0.71-1.80) | 0.585 |
| Type of COVID-19 services delivered |  |  |  |  |
| COVID-19 ward or Intensive care unit (ICU) (n=524) | 0.537 | 0.146 | 1.71(1.28-2.27) | <0.001 |
| Outpatient COVID-19 care (n=490) | -0.237 | 0.150 | 0.78(0.58-1.05) | 0.114 |
| Telemedicine or other distance-based COVID-19 services (n=178) | 0.056 | 0.196 | 1.05(0.72-1.55) | 0.776 |
| Research on COVID-19 (n=61) | -0.226 | 0.312 | 0.79(0.43-1.47) | 0.470 |
| Epidemiological surveillance or COVID-19 boards (n=53) | 0.118 | 0.333 | 1.12(0.58-2.16) | 0.723 |

At the time of administration of the survey, the vast majority of physicians declared having already been vaccinated (93%), with non-significant differences in the vaccine uptake among Maranhão and São Paulo physicians. 35.8% of all the physicians declared having been infected with COVID-19 in the previous year. Almost half (49.2%) [CI 45.0-53.4] of Maranhão physicians were infected, with the majority of them having suffered only mild or no symptoms (42.3%) (Fig.1). Conversely, in São Paulo 24.1% of physicians [CI 20.8 - 27.5] declared a COVID-19 infection, again with the majority recalling mild or no symptoms (Fig. 1). In our sample, 4.5% of physicians were affected by severe COVID-19 symptoms, with the proportion in Maranhão (6.9%) [CI 5.0 - 9.2] significantly higher than in São Paulo (2.5%) [CI 1.5 - 4.0].

**Figure 1:**
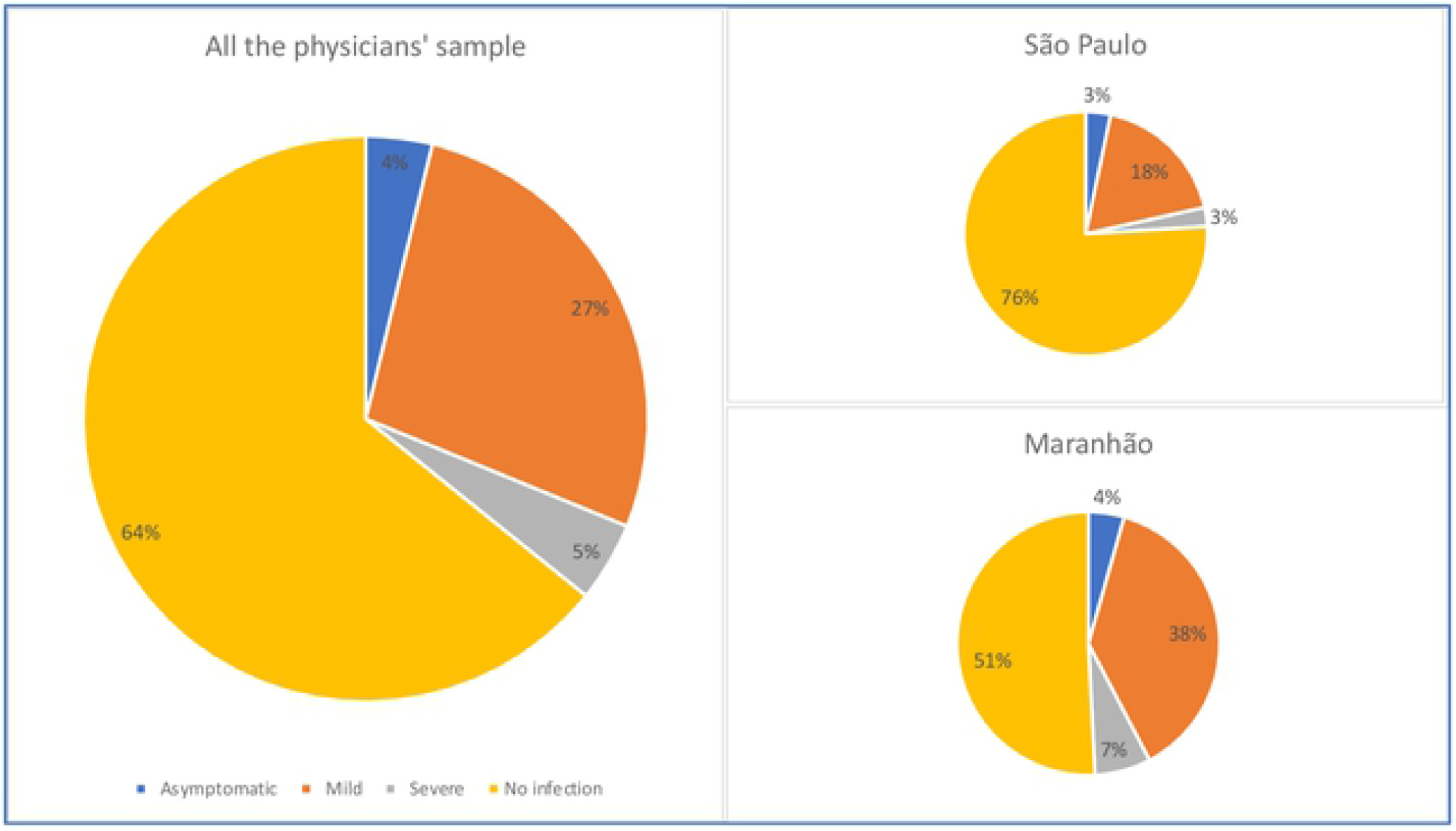
Physician infection rates in the sample locations.

Such infections directly resulted in the loss of 7,117 workdays, representing 2.36% of the total working days in our physician sample.

The multivariate analysis showed that being from Maranhão State [95% CI 2.08-3.57], younger than 50 years [95% CI 1.41-2.89], and having worked in a COVID-19 ward [95% CI 1.28-2.27], were positively associated with the odds of contracting COVID-19 (Tab. 3). Conversely, working with diagnostic services such as X-rays or scans [95% CI 0.53-0.96], in administrative functions [95% CI 0.42-0.80], or in teaching and research [95% CI 0.48-0.91] had a protective effect on the probability of contracting COVID-19.

When multivariate models were run separately for each state, the results were not materially different, except for ‘Working in diagnostic services’ that was no longer significant for the São Paulo cohort (see Supporting information 3: Maranhão and São Paulo multivariate regression models).

With specific reference to ‘Age’, when using ‘>50 years’ as reference category, both the ‘35-50 years’ and ‘35-50 year’ categories showed strong, significant association with the outcome variable of likelihood of COVID-19 infections, although the latter appeared to have higher significance (p<0.001) and coefficient (B=0.706).

On the association between physicians’ engagement in specific services and probability of getting infected, only being deployed in COVID-19 hospital wards or ICU displayed a strong positive association for the two states, while deployment in administrative services displayed a strong negative significance. The significance of being deployed in diagnostic services was only significant for the Maranhão cohort, and deployment in teaching and research activities for the São Paulo cohort, loosing significance when analysed jointly across the sample.

## 4 Discussion

Our survey found that 35.8% of physicians in two Brazilian states were infected with COVID-19 in the first year of the pandemic. Although most of them only experienced mild symptoms, 7,117 workdays were lost following the infections. The rate of infections varied considerably between Maranhão and São Paulo physicians, with the former being affected twice as much than the latter. Being a Maranhão physician, being younger than 50 years old, and deployed to a COVID-19 ward, were the factors positively associated with infections. Conversely, being deployed to diagnostic services, administrative functions, or to teaching and research, were found to have a protective effect. Physician gender, sector of employment, and deployment to other frontline services, were not significantly associated with infections.

These findings need to be interpreted with a degree of caution. Our survey sample did not include those physicians who died or were critically incapacitated because of COVID-19, therefore possibly underestimating overall infections. Our data on infections, personal and professional characteristics were based on physicians’ responses, which could have been affected by recall bias (33). We did not collect information on availability and use of Personal Protective Equipment (PPE) among our physicians, which is considered in the COVID-19 literature a key risk factor for infections.(14) Finally, Maranhão and São Paulo represent very particular settings in terms of income distribution, organisation of healthcare services, and labour markets characteristics;(34) therefore, the study findings may not be entirely generalisable to the rest of Brazil, let alone to other low- and middle-income countries. Despite these limitations, a few conclusions can be safely drawn from our work.

We show that over a third of the medical workforce in Maranhão and São Paulo was infected with COVID-19 in the first year of the pandemic, with a substantial loss of labour. This is consistent with the findings from smaller studies from Brazil(20) and other LMICs,(16,18,19) and therefore particularly relevant for those countries with a scarcity of healthcare resources, which will have been hit already particularly hard by the pandemic (35).

The higher infection rate among Maranhão physicians was in contrast to lower population infection rates (see Tab.1). Our multivariate analysis confirmed that working in Maranhão was one of the most significant risk factors of physician infections in our cohort. The lower ratio of physicians per capita in Maranhão (1.1 per 1,000 in Maranhão Vs 3.2 per 1,000 in São Paulo)(27) may be a factor here, as during health emergencies a smaller workforce will necessarily engage in multiple functions and tasks across sectors, therefore increasing opportunities for infection. This is consistent with previous work(26) showing the differential impact of health system crises on unequal states in LMICs. If confirmed, such finding would be relevant for those studies forecasting effects of the pandemic on health workforces in different parts of the world (4)

Younger age was associated with higher infection rates among physicians in both Brazilian states, which on the one hand contradicts the existing evidence on COVID-19 risk factors from high-income countries,(36) but on the other, is consistent with the scant evidence from other LMICs.(16–19) New public and private medical schools are supplying the national workforce(37), newly graduated physicians were predominantly deployed to COVID-19 wards and Nightingale hospitals, as older physicians were excluded from COVID-19 functions because considered at risk.(38)(39) While such a policy may make sense from a health management and epidemiological point of view, it also poses ethical and equity questions on medical employment in LMICs,(40)(41) particularly at a time when contracts of newly graduated physicians suffer for increasing casualisation in Brazil and worldwide.(42)(43)

Not unexpectedly, our analysis also shows that working in COVID-19 wards is associated with higher rates of infections among physicians compared to working in administrative functions, teaching and research. Although this is consistent with what is already known on COVID-19 risk factors for health workers(9,14), it is however surprising that other ‘frontline’ functions (such as COVID-19 outpatient visits or diagnostic services) were not also significantly associated with increased odds of infections. Such a lack of effect could be of course explained by the suspension of some ‘non-essential’ services during the pandemic, which would have spared physicians from dangerous exposure (44). But it is also likely that the definition of what constitute ‘frontline health services’ may not be that clear-cut, particularly in LMICs settings during an epidemic, where boundaries and functions become blurred, and health workers end up carrying out whatever functions are needed.

It was also surprising to see similar odds of infections among public and private sectors physicians, particularly as publicly funded health systems are believed to have borne the brunt of the COVID-19 pandemic (45,46), particularly in Brazil.(47) In the case of Brazilian’s physicians it is more likely that blurred boundaries between public and private employment may have made it difficult to accurately distinguish between physicians of the Unified National Healthcare System (SUS), from those of the private sector. As previous research has shown that the majority of Brazilian doctors simultaneously engage in a multiplicity of public as well as private forms of employment (48), and that private organisations often provide services within SUS (34), it is likely that the majority of physicians in our sample carried out functions simultaneously in public and private sector institutions, making it very difficult to identify the individual effects of the pandemic on either sectors.

If confirmed, the findings from this study have policy implications for the ongoing efforts to estimate the effects of the pandemic on HWs, as well as for health policymakers in LMICs. Emerging evidence from different countries hints that the WHO’s high-end figure(4) of 180,000 COVID-19 deaths among HWs is probably an overestimation. Rather than applying population-based infection and mortality rates, our study suggests that rates among HWs may vary according to HWs density, with understaffed health systems and services suffering more infections and deaths. This could help refine projections, and re-focus policies to mitigate the impact of the pandemic on health workforces worldwide.

Although medical doctors might not be the HWs most at risk of COVID-19 infections,(10) our study suggests that COVID-19’s impact on such key and expensive human resource is not negligible, and less staffed parts of a country’s health systems may be more at risk. Health authorities worldwide should therefore give priority during epidemics to protect those services with the lowest HW to population ratios, knowing that opportunities for infections will be greater where workers are more scarce.(45)

Finally, our work appears to confirm that younger doctors in LMICs may be more at risk of infection than their counterparts in HICs,(17,19) possibly because the health services in those countries have drawn from younger recruits to staff COVID-19 services. While such decisions can be justified in the light of younger doctors’ less severe hospitalization and fatality rates, health authorities should make sure younger doctors are compensated through better contracts, and access to more secure parts of the market for physician services (49).

## 5 Conclusions

Health workers are essential resources for any health system, and they have borne the brunt of providing life-saving services during the COVID-19 epidemic. Knowledge gaps exist on COVID-19 infections, mortality and risk factors among HWs, particularly in LMICs. We conducted a cross-sectional telephone survey in a representative sample of the physicians in the populous São Paulo, and the disadvantaged Maranhão state in Brazil, with a view to identify the associated risk factors for such a key medical profession.

We found that more than a third of physicians in two states were infected with COVID-19 in the first year of the pandemic. Although most of them only experienced mild symptoms, a substantial number of workdays were lost following the infections. The rate of infections varied considerably between Maranhão and São Paulo physicians, with the former being affected twice as much than the latter. Being a Maranhão physician, younger than 50 years, and deployed to a COVID-19 ward, were the factors positively associated with infections. Conversely, being deployed to diagnostic services, administrative functions, or to teaching and research, were found to have a protective effect.

More research is needed to explore depth and nuances of the impact of the pandemic on HWs in LMICs. Our findings on the greater impact for physicians from less-staffed parts of Brazil’s health system carry implications for the identification of policy to mitigate COVID-19 effects on health workforces, and for their measurement worldwide.

## Data Availability

The survey dataset has now been attached to this submission - see the Supporting information material 'DBase survey physicians in Brazil'.

## Acknowledgements and funding

We are indebted to Prof. Trevor Sheldon and Dr. James Buchan for their valuable comments and suggestions to an early version of the manuscript.

This study received support from the Confap-MRC call for Health Systems Research Networks. Specifically, GR received the award from the Newton Fund/ Medical Research Council (UK), Grant Reference MR/R022747/1. AJC and MS received the award from the *Fundação de Amparo à Pesquisa do Estado de São Paulo* (FAPESP-Brazil), 2017/50356-7. BdO received the award from the *Fundação de Amparo à Pesquisa e ao Desenvolvimento Científico e Tecnológico do Maranhão* (FAPEMA-Brazil), COOPI-00709/18.

## Contributor statement

GR designed the study, conducted the analysis, and drafted the manuscript. AJC conducted the analysis, and contributed to data collection. BdO conducted the analysis, and participated in the data collection. MS designed the study, and revised the manuscript. All the authors read and approved the final version of the manuscript.

## Supporting information captions

Supporting information 1: Sampling equations for replacement

Supporting information 2: Survey questionnaire

Supporting information 3: Maranhão and São Paulo regression models

## References

1. Campbell J, Dussault G, Buchan J, Pozo-Martin F, Cometto G. A Universal Truth: No Health Without a Workforce [Internet]. Recife, Brazil: Global Health Workforce Alliance; 2013 [cited 2022 Mar 8]. Available from: https://www.who.int/workforcealliance/knowledge/resources/hrhreport2013/en/

2. Mehta S, Machado F, Kwizera A, Papazian L, Moss M, Azoulay É, et al. COVID-19: a heavy toll on health-care workers. The Lancet Respiratory Medicine. 2021 Mar 1;9(3):226–8.

3. The Lancet. COVID-19: protecting health-care workers. Lancet. 2020;395(10228):922.

4. WHO. The impact of COVID-19 on health and care workers: a closer look at deaths [Internet]. Geneva, Switzerland: Health Workforce Department; 2021. Report No.: WHO/HWF/WorkingPaper/2021.1. Available from: https://apps.who.int/iris/bitstream/handle/10665/345300/WHO-HWF-WorkingPaper-2021.1-eng.pdf?sequence=1&isAllowed=y

5. Vargese SS, Dev SS, Soman A S, Kurian N, Varghese V A, Mathew E. Exposure risk and COVID-19 infection among frontline health-care workers: A single tertiary care centre experience. Clinical Epidemiology and Global Health. 2022 Jan 1;13:100933.

6. Iversen K, Bundgaard H, Hasselbalch RB, Kristensen JH, Nielsen PB, Pries-Heje M, et al. Risk of COVID-19 in health-care workers in Denmark: an observational cohort study. The Lancet Infectious Diseases. 2020 Dec 1;20(12):1401–8.

7. Dzinamarira T, Murewanhema G, Mhango M, Iradukunda PG, Chitungo I, Mashora M, et al. COVID-19 Prevalence among Healthcare Workers. A Systematic Review and Meta-Analysis. Int J Environ Res Public Health. 2021 Dec 23;19(1):146.

8. Bandyopadhyay S, Baticulon RE, Kadhum M, Alser M, Ojuka DK, Badereddin Y, et al. Infection and mortality of healthcare workers worldwide from COVID-19: a systematic review. BMJ Global Health. 2020 Dec 1;5(12):e003097.

9. Chou R, Dana T, Buckley DI, Selph S, Fu R, Totten AM. Epidemiology of and Risk Factors for Coronavirus Infection in Health Care Workers. Ann Intern Med. 2020 Jul 21;173(2):120–36.

10. Gómez-Ochoa SA, Franco OH, Rojas LZ, Raguindin PF, Roa-Díaz ZM, Wyssmann BM, et al. COVID-19 in Healthcare Workers: A Living Systematic Review and Meta-analysis of Prevalence, Risk Factors, Clinical Characteristics, and Outcomes. Am J Epidemiol. 2020 Sep 1;kwaa191.

11. Chou R, Dana T, Buckley DI, Selph S, Fu R, Totten AM. Update Alert 10: Epidemiology of and Risk Factors for Coronavirus Infection in Health Care Workers. Ann Intern Med. 2022 Jan 18;175(1):W8–9.

12. Gholami M, Fawad I, Shadan S, Rowaiee R, Ghanem H, Hassan Khamis A, et al. COVID-19 and healthcare workers: A systematic review and meta-analysis. International Journal of Infectious Diseases. 2021 Mar 1;104:335–46.

13. Shah ASV, Wood R, Gribben C, Caldwell D, Bishop J, Weir A, et al. Risk of hospital admission with coronavirus disease 2019 in healthcare workers and their households: nationwide linkage cohort study. BMJ. 2020 Oct 28;371:m3582.

14. Nguyen LH, Drew DA, Graham MS, Joshi AD, Guo CG, Ma W, et al. Risk of COVID-19 among front-line health-care workers and the general community: a prospective cohort study. The Lancet Public Health. 2020 Sep 1;5(9):e475–83.

15. Shields A, Faustini SE, Perez-Toledo M, Jossi S, Aldera E, Allen JD, et al. SARS-CoV-2 seroprevalence and asymptomatic viral carriage in healthcare workers: a cross-sectional study. Thorax. 2020 Dec 1;75(12):1089–94.

16. Liu J, Ouyang L, Yang D, Han X, Cao Y, Alwalid O, et al. Epidemiological, Clinical, Radiological Characteristics and Outcomes of Medical Staff with COVID-19 in Wuhan, China: Analysis of 101 Cases. Int J Med Sci. 2021 Jan 29;18(6):1492–501.

17. Sabetian G, Moghadami M, Hashemizadeh Fard Haghighi L, Shahriarirad R, Fallahi MJ, Asmarian N, et al. COVID-19 infection among healthcare workers: a cross-sectional study in southwest Iran. Virol J. 2021 Mar 17;18(1):58.

18. Alajmi J, Jeremijenko AM, Abraham JC, Alishaq M, Concepcion EG, Butt AA, et al. COVID-19 infection among healthcare workers in a national healthcare system: The Qatar experience. International Journal of Infectious Diseases. 2020 Nov 1;100:386–9.

19. Al Abri ZGH, Al Zeedi MASA, Al Lawati AA. Risk Factors Associated with COVID-19 Infected Healthcare Workers in Muscat Governorate, Oman. J Prim Care Community Health. 2021 Jan 1;12:2150132721995454.

20. Correia RF, Costa ACC da, Moore DCBC, Junior SCG, Oliveira MPC de, Zuma MCC, et al. SARS-CoV-2 seroprevalence and social inequalities in different subgroups of healthcare workers in Rio de Janeiro, Brazil. The Lancet Regional Health – Americas [Internet]. 2022 Mar 1 [cited 2022 Feb 15];7. Available from: https://www.thelancet.com/journals/lanam/article/PIIS2667-193X(21)00166-6/fulltext

21. Oliveira MS de, Lobo RD, Detta FP, Vieira-Junior JM, Castro TL de S, Zambelli DB, et al. SARS-Cov-2 seroprevalence and risk factors among health care workers: Estimating the risk of COVID-19 dedicated units. American Journal of Infection Control. 2021 Sep 1;49(9):1197–9.

22. Buonafine CP, Paiatto BNM, Leal FB, de Matos SF, de Morais CO, Guerra GG, et al. High prevalence of SARS-CoV-2 infection among symptomatic healthcare workers in a large university tertiary hospital in São Paulo, Brazil. BMC Infectious Diseases. 2020 Dec 2;20(1):917.

23. Ritchie H, Mathieu E, Rodés-Guirao L, Appel C, Giattino C, Ortiz-Ospina E, et al. Coronavirus Pandemic (COVID-19). Our World in Data [Internet]. 2020 Mar 5 [cited 2022 Mar 9]; Available from: https://ourworldindata.org/coronavirus-source-data

24. UKRI. How is the current crisis reshaping Brazil’s health system? Strengthening health workforce and provision of services in São Paulo and Maranhão [Internet]. UKRI gateway to publicly funded research and innovation. 2018 [cited 2022 Mar 9]. Available from: https://gtr.ukri.org/projects?ref=MR%2FR022747%2F1&pn=0&fetchSize=10&selectedSortableField=title&selectedSortOrder=ASC

25. Paim J, Travassos C, Almeida C, Bahia L, Macinko J. The Brazilian health system: history, advances, and challenges. The Lancet. 2011 May 21;377(9779):1778–97.

26. Andrietta LS, Levi ML, Scheffer MC, Alves MTSS de B e, Oliveira BLCA de, Russo G. The differential impact of economic recessions on health systems in middle-income settings: a comparative case study of unequal states in Brazil. BMJ Global Health. 2020 Feb 1;5(2):e002122.

27. Scheffer MC, Cassenote A, Guilloux AGA, Guerra A, Miotto BA. Demografia médica no Brasil 2020 [Internet]. São Paulo (BRA): Faculdade de Medicina USP, Conselho Federal de Medicina; 2021 p. 125. Available from: https://www.fm.usp.br/fmusp/conteudo/DemografiaMedica2020_9DEZ.pdf

28. CONASS. PAINEL CONASS | COVID-19 [Internet]. 2022 [cited 2022 Mar 14]. Available from: https://www.conass.org.br/painelconasscovid19/

29. Burgess S, CRP CHD Genetics Collaboration. Identifying the odds ratio estimated by a two-stage instrumental variable analysis with a logistic regression model. Stat Med. 2013 Nov 30;32(27):4726–47.

30. Hamaker EL, Hattum P van, Kuiper RM, Hoijtink H. Model Selection Based on Information Criteria in Multilevel Modeling. In: Handbook of Advanced Multilevel Analysis. Routledge; 2010.

31. Park S, Lake ET. Multilevel modeling of a clustered continuous outcome: nurses’ work hours and burnout. Nurs Res. 2005 Dec;54(6):406–13.

32. R Core Team — European Environment Agency. R: A language and environment for statistical computing [Internet]. 2020 [cited 2022 Mar 9]. Available from: https://www.eea.europa.eu/data-and-maps/indicators/oxygen-consuming-substances-in-rivers/r-development-core-team-2006

33. Althubaiti A. Information bias in health research: definition, pitfalls, and adjustment methods. J Multidiscip Healthc. 2016 May 4;9:211–7.

34. Russo G, Levi ML, de Britto e Alves MTSS, de Oliveira BLCA, de Souza Britto Ferreira de Carvalho RH, Andrietta LS, et al. How the ‘plates’ of a health system can shift, change and adjust during economic recessions. A qualitative interview study of public and private health providers in Brazil’s São Paulo and Maranhão states [Internet]. SocArXiv; 2020 Jul [cited 2020 Jul 14]. Available from: https://osf.io/qtgux

35. Blanchet K, Alwan A, Antoine C, Cros MJ, Feroz F, Guracha TA, et al. Protecting essential health services in low-income and middle-income countries and humanitarian settings while responding to the COVID-19 pandemic. BMJ Global Health. 2020 Oct 1;5(10):e003675.

36. Ho FK, Petermann-Rocha F, Gray SR, Jani BD, Katikireddi SV, Niedzwiedz CL, et al. Is older age associated with COVID-19 mortality in the absence of other risk factors? General population cohort study of 470,034 participants. PLOS ONE. 2020 Nov 5;15(11):e0241824.

37. Scheffer MC, Pastor-Valero M, Cassenote AJF, Compañ Rosique AF. How many and which physicians? A comparative study of the evolution of the supply of physicians and specialist training in Brazil and Spain. Hum Resour Health. 2020 Apr 21;18(1):30.

38. Sodré F. Epidemia de Covid-19: questões críticas para a gestão da saúde pública no Brasil. Trab educ saúde [Internet]. 2020 Aug 28 [cited 2022 Mar 11];18. Available from: http://www.scielo.br/j/tes/a/YtCRHxTywqWm4SChBHvqPBB/?lang=pt

39. Inácio AM. Covid-19. Médicos com mais de 60 anos são grupo de risco. Devem ou não ser poupados? [Internet]. [cited 2022 Mar 31]. Available from: https://www.dn.pt/pais/covid-19-medicos-com-mais-de-60-anos-sao-grupo-de-risco-devem-ou-nao-ser-poupados-11962500.html

40. Scheffler RM. The labour market for human resources for health in low-and middle-income countries. Geneva; 12 p.

41. McPake B, Scott A, Edoka I. Analyzing Markets for Health Workers: Insights from Labor and Health Economics. World Bank Publications; 2014. 99 p.

42. Econômico B. STF libera pejotização de médicos e especialistas alertam para riscos [Internet]. iG. 2022 [cited 2022 Mar 24]. Available from: https://economia.ig.com.br/2022-02-18/stf-libera-pejotizacao-de-medicos-e-especialistas-alertam-para-riscos.html

43. ILO. Inequalities and the world of work [Internet]. Geneva, Switzerland: International Labour Organization; 2021 [cited 2022 Mar 24]. Report No.: ISBN 978-92-2-034165-0. Available from: <http://www.ilo.org/ilc/ILCSessions/109/reports/reports-to-the-conference/WCMS_792123/lang--en/index.htm>

44. Russo G, Jesus TS, Deane K, Osman AY, McCoy D. Epidemics, Lockdown Measures and Vulnerable Populations: A Mixed-Methods Systematic Review of the Evidence of Impacts on Mother and Child Health in Low-And-Lower Middle-Income Countries. Int J Health Policy Manag. 2021;1.

45. Haldane V, De Foo C, Abdalla SM, Jung AS, Tan M, Wu S, et al. Health systems resilience in managing the COVID-19 pandemic: lessons from 28 countries. Nat Med. 2021 Jun;27(6):964–80.

46. Tessema GA, Kinfu Y, Dachew BA, Tesema AG, Assefa Y, Alene KA, et al. The COVID-19 pandemic and healthcare systems in Africa: a scoping review of preparedness, impact and response. BMJ Global Health. 2021 Dec 1;6(12):e007179.

47. Dal Poz MR, Levcovitz E, Bahia L. Brazil’s Fight Against COVID-19. Am J Public Health. 2021 Mar;111(3):390–1.

48. Miotto BA, Guilloux AGA, Cassenote AJF, Mainardi GM, Russo G, Scheffer MC. Physician’s sociodemographic profile and distribution across public and private health care: an insight into physicians’ dual practice in Brazil. BMC Health Services Research. 2018 Apr 23;18:299.

49. Mcpake B, Squires AP, Mahat A, Araujo EC. The economics of health professional education and careers: insights from a literature review [Internet]. The World Bank; 2015 Sep [cited 2018 Feb 5] p. 1–89. Report No.: 99535. Available from: http://documents.worldbank.org/curated/en/570681468190783192/The-economics-of-health-professional-education-and-careers-insights-from-a-literature-review

